# An integrative study of risk assessment, mediation analysis, and causal inference for the relationship between metabolic syndrome and dilated cardiomyopathy

**DOI:** 10.64898/2026.04.07.26350282

**Authors:** Jike Qi, Shuo Zhang, Yuchen Jiang, Hua Lin, Chu Zheng, Yu Yan, Ke Wang, Hongyan Cao, Weiyi Song, Yue Xu, Ping Zeng

**Author notes:** Co-first authors (Jike Qi, Shuo Zhang, Yuchen Jiang). **Correspondence to:** Ping Zeng and Yue Xu, Department of Biostatistics and Jiangsu Engineering Research Center of Biological Data Mining and Healthcare Transformation, Xuzhou Medical University, Xuzhou, Jiangsu 221004, China.; or.

## Abstract

**Aims:** While metabolic syndrome (MetS) is observed in dilated cardiomyopathy (DCM) patients, its causal role in disease development remains unestablished. This study aims to assess if MetS is a causative risk factor for DCM incidence.

**Materials and methods:** By leveraging 378,837 UK Biobank participants, instead of the conventional binary MetS, we calculated a continuous metabolic risk score (MRS) and evaluated its influence on DCM risk within a multi-model evidence framework. Bidirectional weighted quantile sum regression identified key MRS components, a nested case-control study assessed 14-year pre-diagnosis MRS trajectories, mediation analyses evaluated MRS mediating lifestyle-DCM links and inflammation mediating MRS-DCM relationships, and Mendelian randomization (MR) evaluated causality for genetically predicted MetS and components on DCM.

**Results:** During a median follow-up period of 13.4 years (interquartile range 12.7∼14.1 years), 820 (0.2%) participants developed DCM. Higher MRS (HR=1.26 [1.18∼1.34]) was associated with increased DCM risk, and such an association persisted across all robustness assessments even among non-MetS individuals. Waist circumference (WC, HR=1.36 [1.28∼1.45], weight=0.58) and glycated hemoglobin (HR=1.23 [1.16∼1.30], weight=0.22) dominated MRS’ risk contribution. The trajectories of MRS diverged in cases approximately 10 years pre-diagnosis. MRS mediated 5.1∼26.2% of lifestyle-related DCM risk, while inflammation mediated 16.4% of the MRS-DCM association. MR analysis further confirmed causal effects of MetS (OR=1.65 [1.45∼1.88]), WC (OR=1.79 [1.58∼2.03]) on DCM risk.

**Conclusions:** Metabolic dysfunction, which was dominated by central adiposity and hyperglycemia, plays a key role in the occurrence of DCM. Early intervention targeting metabolic factors may prevent DCM onset.

**Lay summary:** This study demonstrates that metabolic dysfunction, specifically central adiposity and hyperglycemia, drives causal risk for DCM. Metabolic abnormalities diverge ∼10 years pre-diagnosis, offering a critical window for intervention. MRS mediates lifestyle-related DCM risk (≤26%) and acts partially via inflammation. Early metabolic monitoring in high-risk individuals and tailored interventions targeting these components or their inflammatory consequences could prevent DCM onset and reduce mortality.

## Introduction

Dilated cardiomyopathy (DCM) is a myocardial disorder, which was characterized by ventricular dilation and impaired systolic function, and is a major contributor to heart failure, life-threatening arrhythmias and sudden cardiac death 1. Current evidence suggests the occurrence of DCM primarily derives from the interplay between genetic susceptibility and environmental triggers. However, emerging evidence implies that metabolic dysfunction maybe an unignorable factor in the pathogenesis of DCM 2. Metabolic syndrome (MetS), defined as a cluster of metabolic abnormalities, serves as a crucial composite indicator for assessing systemic metabolic status 3,4. Accumulating epidemiological and clinical evidence has confirmed that MetS is strongly associated with incident major cardiovascular events 3,4.

Research on MetS in DCM has primarily focused on its role after disease onset, revealing complex associations with patient outcomes. One single-center analysis suggested that, counterintuitively, the presence of MetS might be associated with improved survival in this specific patient cohort 5. Concurrently, advanced imaging studies have provided mechanistic insights, demonstrating that MetS can exacerbate adverse cardiac remodeling and impair myocardial efficiency in these patients 6. These correlative findings nevertheless establish a clear association between MetS and DCM pathology. This prompts a critical question: does this relationship begin before diagnosis? Investigating whether MetS is a predisposing risk factor for DCM incidence is essential to trace this link back to its origin and explore preventive potential.

Additionally, in prior work, MetS was predominantly used as a binary indicator (MetS and non-MetS) 7-9, which limits an explicit understanding of the condition. This usage leads to critical drawbacks, including failing to capture the continuous spectrum of metabolic risk, obscuring severity assessment, and excluding individuals with poor metabolic status who fail to reach the diagnostic threshold yet but remain at heightened metabolic risk. Whereas metabolic risk score (MRS), a continuous indicator, is essential to overcome these limitations by quantifying the potential dose-response relationship of metabolic abnormalities 10, enabling a more precise exploration of the MetS-DCM link.

Beyond the shortage of usage of MetS, current methodologies for examining cardiometabolic measurements, including MetS, are also hindered by other limitations. First, previous studies often only focus on composite MetS measures over examining the specific contributions of individual components 7-9, limiting understanding of their unique effects. Second, the absence of long-term follow-up data hampered the characterization of factors’ trajectories leading to outcome onset 11. Third, lifestyle drivers like dietary patterns and alcohol intake, which were known to influence both MetS and DCM 12,13 were typically relegated to mere confounders, thereby undervaluing their pivotal roles in effect mediation. Similarly, the central mediating function of inflammation in many cardiac comorbidities was also frequently sidelined in association analysis, despite its clinical importance 14. Finally, the traditional observational models relied on by most studies are prone to residual confounding and reverse causality, fundamentally limiting the ability to establish reliable causal inference.

To address these limitations, in this work we constructed a multi-model evidence framework to elucidate the relationship between MetS and DCM. First, we calculated MRS in the large-scale UK Biobank cohort and evaluated its association with incident DCM risk. For comparison, we especially analyzed subgroups free of MetS or any adverse metabolic components to better discover subclinical risk. Then, we employed weighted quantile sum regression (WQS) to decompose component-specific weights for MRS. Meanwhile, a nested case-control study tracked longitudinal MRS trajectories in DCM patients, and mediation analysis was conducted to dissect MRS-centered mediation of lifestyle effects on DCM and inflammation cascade mediation linking MRS to DCM pathogenesis. Finally, Mendelian randomization (MR) assessed causal effects of genetically predicted MetS on DCM incidence. This integrative analytical framework is presented in Figure 1.

**Figure 1.**
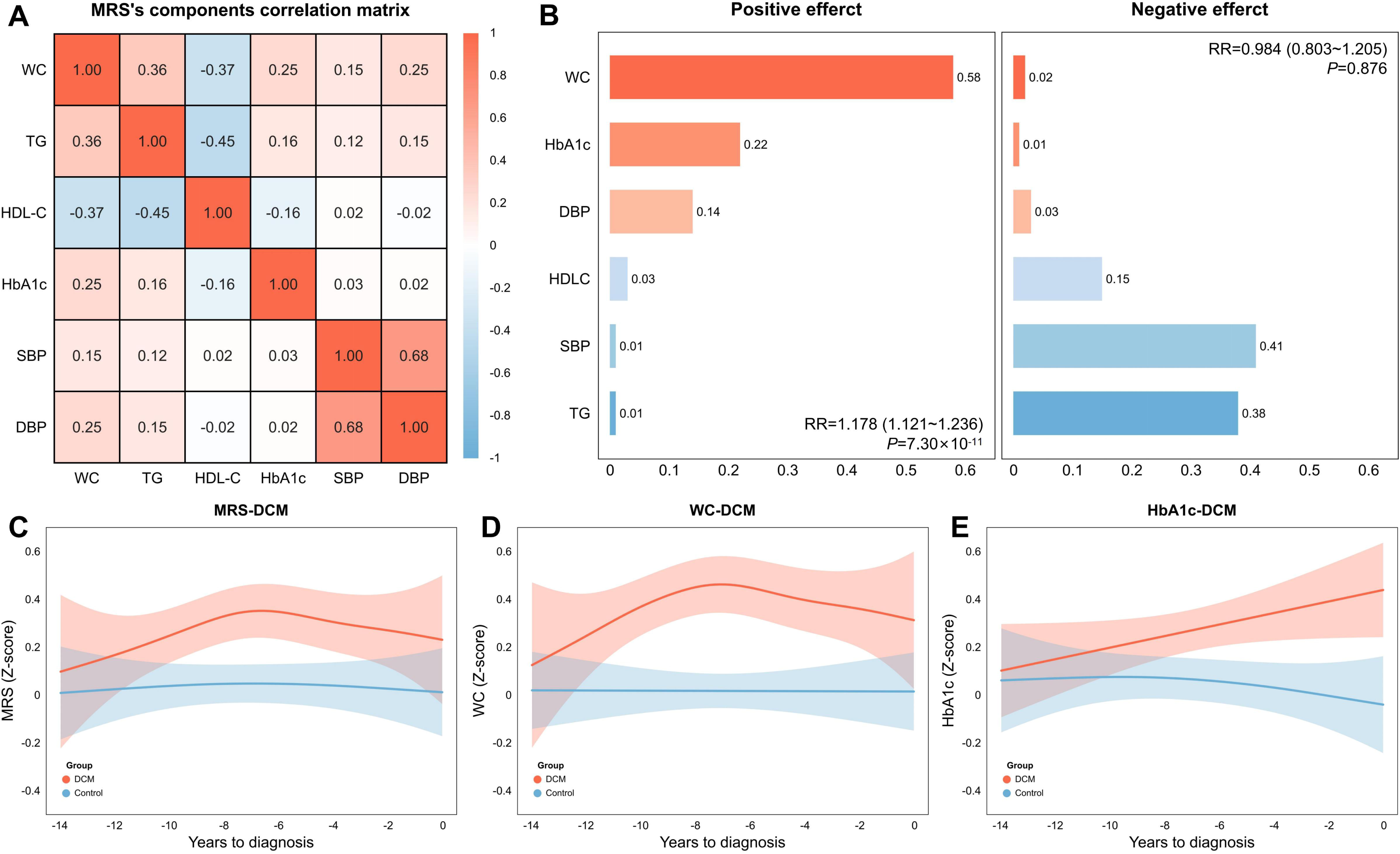
Flow chart of the study. MRS-related traits serve as the exposure, with DCM as the outcome. We investigated the phenotypic and genetic associations between the two. MRS, metabolic risk score; DCM, dilated cardiomyopathy; WC, waist circumference; TG, triglycerides; HDL-C, high-density lipoprotein cholesterol; HbA1c, glycated hemoglobin; BP, blood pressure; PC1, first principal component; GPS, generalized propensity score; WQS, weighted quantile sum regression; MR, Mendelian randomization; IVs, instrumental variables; PSM, propensity score matching; INFLA, low-grade chronic inflammation; *N*, sample size.

## Methods

### Data sources and variable definitions

#### UK Biobank cohort

The UK Biobank 15 is a large, population-based prospective cohort study initiated in 2006-2010. It enrolled over 500,000 participants aged 37-73 years across 22 assessment centers in England, Wales, and Scotland. At baseline, data collection comprised touch-screen questionnaires, nurse-led interviews, physical examinations, and biological sample provision to obtain sociodemographic, lifestyle, environmental, and health-related information. To mitigate potential confounding from population structure and enhance analytical robustness, the present analysis was restricted to participants of white European ancestry. Participants were excluded if they: (i) lacked data necessary to calculate any component of the MRS; these components were waist circumference (WC; field 48), triglycerides (TG; field 30870), high-density lipoprotein cholesterol (HDL-C; field 30760), glycated hemoglobin (HbA1c; field 30750), systolic blood pressure (SBP; fields 93 and 4080) and diastolic blood pressure (DBP; fields 94 and 4079); (ii) had HbA1c values in the top 1% of the distribution to exclude extreme outliers; (iii) had a baseline diagnosis of DCM (field 40720-I42.0); or (iv) were lost to follow-up or withdrew consent.

#### Definition of the MRS and DCM

Referring to previous studies 10,16, we constructed MRS via

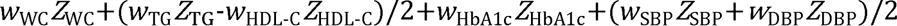

where *Z* represents age- and sex-standardized residuals and *w* denotes weights for each component. SBP and DBP were additionally standardized for height to eliminate the effect of height, and TG was log-transformed to handle the skewed distribution. We set equal weights for all the components. A higher MRS indicates more severe metabolic disorders. The follow-up of each participant was defined as the time span from baseline to the onset of DCM, death, or censoring date (July 19, 2022), whichever came first. DCM cases were defined on the basis of the ICD-10.

#### Selection of covariates

Considering age, sex, and height were already adjusted in the MRS calculation, these covariates were employed only for secondary exposures and PSM. In addition, we selected socioeconomic factors (Townsend deprivation index [TDI], educational level, and income), lifestyle factors (alcohol consumption history, smoking history, physical activity status, and healthy diet score), and the top 10 genetic principal components (PC) at baseline as covariates (Method S1). Multiple imputation by chained equations was used to estimate missing values (Method S1) for each covariate 17.

#### GWAS summary statistics

Given the absence of genome-wide association studies specifically investigating MRS, we utilized the largest available summary statistics of genome-wide association studies (GWAS) for MetS (*N*=1,384,348) as a genetically correlated proxy 18. Summary-level data for DCM (*N*_case_=14,256; *N*_control_=1,199,156) were extracted from a meta-analysis of 16 studies 19. Details on the summary statistics for MetS, its significant components and DCM are provided in Table S1. Rigorous quality control procedures were applied to each summary statistics dataset (Method S2).

### Statistical analyses

#### Survival analyses evaluating the relationship between MRS and DCM

Our observational analysis was implemented under the STROBE guidelines 20, with checklists shown in Table S2. Baseline characteristics are reported as the mean ± standard deviation (SD) for continuous variables and as the frequency (proportion) for categorical variables. These variables were compared with *t* tests or χ^2^ tests, as appropriate. Multivariate Cox proportional hazards models were employed to evaluate the influence of the MRS on incident DCM, adjusting for selected covariates. The proportional hazards assumption was verified through Schoenfeld’s residual test (global *P*>0.05) 21. The results are presented as hazard ratios (HRs) with 95% confidence intervals (CIs). Restricted cubic spline analysis with 3 knots was used to evaluate potential nonlinear associations between the MRS and DCM risk. The participants were stratified into quartiles (Q1∼Q4) on the basis of the MRS, with Q1 as the reference, to examine dose-response relationships.

As a complementary and comparative analysis for MRS, we further defined binary MetS 22 and included it, along with its five individual components and their cumulative count, as secondary exposures (Method S3). To explore whether the MRS could also affect DCM risk among individuals with healthy metabolic status, highlighting its potential as a broad-spectrum indicator, we restricted the MRS analysis to non-MetS individuals and examined the association specifically in participants without any adverse MetS components.

#### Sensitivity and subgroup analyses

##### Expanding dimensions of MRS

To assess the stability of MRSs calculated with equal weights, we further examined MRSs constructed with positive WQS weights 23, normalized Cox effect sizes, and first PC loadings (PC1) (Method S4). As the first two weighting approaches were outcome-supervised, we thus conducted Monte Carlo cross-validation (1,000 iterations via 50%:50% training-validation splits) 24: the component weights derived from the training set were used to reconstruct the weighted MRS, which was then tested for its influence on DCM risk in the validation set. In contrast, PC1-based weights, which represent an unsupervised method, were computed via the full dataset.

##### Assessment of hidden biases

To assess the relationship between the MRS and DCM robustly, we conducted a series of sensitivity analyses. First, to reduce potential bias from reverse causation, participants who developed DCM or died within the first 2 years of follow-up were excluded. Second, the Fine-Gray model 25 was employed to address competing risks directly from non-DCM-related deaths, thereby mitigating potential distortions inherent in standard Cox proportional hazards estimates. Third, generalized propensity scores (GPSs) 26 were estimated for MRSs as a continuous exposure; here, covariate balance was assessed via standardized mean differences. Finally, to assess the resilience of the observed HRs to unmeasured confounding, we calculated E-values 27, which quantify the minimum strength of an unmeasured confounder required to invalidate the observed HRs.

##### Stratification by sex, age, and age of DCM onset

Recognizing the biological specificity of age and sex, subgroup analyses were conducted by stratifying participants by sex (male and female) and median age (<58.6 and ≥58.6 years). To assess temporal differences in risk, incident DCM cases were grouped by median age at onset (<7.25 and ≥7.25 years) from baseline, and separate Cox models were fitted in each group.

#### Contributions of individual MRS components to MRS-DCM associations

To evaluate the relative contributions of individual MRS components within the composite MRS to DCM risk, we WQS employed regression 23. Bidirectional WQS models were fitted, one was used to identify risk-increasing factors and the other to identify potential protective factors. The full dataset was randomly divided into a training subset (60%) for estimating component weights and a validation subset (40%) for evaluation. Weight estimation utilized 1,000 bootstrap replications within the training set to assess the stability of the weights. The relative risk (RR) along with 95%CI was derived to quantify associations between the WQS index and DCM incidence.

#### Nested case-control study *assessing* time-dependent metabolic shifts preceding DCM diagnosis

To map the dynamic trajectories of the MRS and its significant components preceding DCM diagnosis, we conducted a nested case-control study 28. We defined an observation window for each case from baseline to their event date, with matched controls assigned the same baseline-to-event-duration period. Within this framework, the MRS and its significant components during the 14-year pre-event period (−14 to 0 years) were analyzed. Dynamic trajectories were modelled with locally weighted scatterplot smoothing, enabling direct comparison of pre-outcome trends within temporally aligned case-control pairs.

#### Mediation of lifestyle-DCM via MRS and MRS-DCM via inflammatory markers

To evaluate the mediating role of MRS between lifestyle and DCM, we performed a mediation analysis 29. We defined a composite healthy lifestyle score and its components (alcohol consumption, smoking history, physical activity status, healthy diet score, sedentary duration, healthy sleep score, social isolation score) as exposure, MRS and its significant components as mediators, and DCM as a time-to-event outcome. Additionally, to investigate whether inflammation mediated the association between MRS and DCM, another mediation analysis was performed with MRS and its significant components as the exposure and the low-grade inflammation score (INFLA) as the mediator 30. Details of the lifestyle score and INFLA are provided in Method S5. Cox models were used to assess the exposure-outcome and mediator-outcome associations, whereas linear regression was used to analyse the exposure-mediator link. The mediated proportion (*P_m_*) and its 95%CI were estimated.

#### Genetic causal analyses examining the risk effect of MetS on DCM

##### Two-sample MR

With carefully selected instrumental variables (IVs; Method S6), we conducted two-sample MR analyses to investigate the causal relationship between genetically predicted MetS and its significant components and DCM, adhering to the STROBE-MR guidelines (Table S3). The phenotypic variance explained (PVE) and the *F*-statistic were calculated for each IV to assess instrument strength 31,32. Six complementary methods were employed: the inverse variance weighted (IVW) method, maximum likelihood method, weighted median method, weighted mode method, constrained maximum likelihood and model averaging (cML-MA) method, and contaminated mixture (ConMix) method (Method S6). The appropriate IVW model (fixed- or random-effects) was selected on the basis of Cochran’s Q statistic.

IVW served as the primary effect estimator, and the remaining five provided robustness checks to reduce false-positive risk. Statistical significance was defined as significance from IVW and at least two other methods with the same effect directions. Suggestive significance required only IVW significance.

##### Sensitivity analyses of MR

To ensure the reliability of the MR findings, we performed sensitivity analyses. Horizontal pleiotropy was assessed with Egger’s intercept test to identify potential confounding pathways unrelated to exposure. Outlier detection employs MR-PRESSO, supplemented by leave-one-out analysis, to verify bias robustness. Additionally, directionality of causality was confirmed via MR Steiger to preclude reverse causality bias.

### Statistical software and packages

All analyses were conducted using R (v4.5.0), and all statistical tests were two-sided with a significance threshold of 0.05. Missing data were imputed via the mice package (v3.17.0). Cox proportional hazards modelling employed survival (v3.4.0) and survminer (v0.4.9). Fine-Gray competing risk analysis utilized cmprsk (v2.2-12), and E-value calculations were performed with EValue (v4.1.3). WQS regression, causal mediation analysis, and PSM were implemented through qwqs (v4.1.0), CMAverse (v0.1.0), and MatchIt (v4.7.2). The MR analyses used TwoSampleMR (v0.6.8), MendelianRandomization (v0.1.0), and MRcML.

## Results

### Baseline characteristics

We analyzed 378,837 participants (54.1% female) with an average age of 57.3 years (SD=8.0) (Table 1). During a median follow-up period of 13.4 years (interquartile range 12.7∼14.1 years), 820 (0.2%) developed DCM. Compared to non-DCM participants, DCM patients were older (60.7 vs. 57.3 years), predominantly male (68.4% vs. 45.9%). Socioeconomically, DCM cases showed higher TDI (−1.1 vs. −1.5) and lower income (e.g., <£18,000: 26.7% vs. 18.5%). Lifestyle factors included elevated smoking rates (66.2% vs. 61.1%) and lower physical activity (e.g., low level physical activity: 19.9% vs. 15.0%), and poorer diet scores (3.0 vs. 3.2) were also observed for DCM patients. Metabolic profiles were consistently adverse in DCM patients, with elevated WC (98.0 vs. 90.1 cm), TG (1.9 vs. 1.7 mmol/L), HbA1c (37.4 vs. 35.5 mmol/mol), and blood pressure (e.g., SBP: 142.2 vs. 140.0 mmHg), alongside reduced HDL-C (1.4 vs. 1.5 mmol/L), translating to higher MRS (0.7 vs. 0.0).

### Estimated effect of the MRS on DCM risk

First, we found that participants meeting the diagnostic criteria demonstrated a 35.5% (17.1∼56.8%) higher DCM risk than did those without MetS. In addition to the expected associations with elevated WC (HR=1.64 [1.43∼1.88]) and elevated HbA1c (HR=1.64 [1.34∼2.01]), reduced HDL-C (HR=1.30 [1.10∼1.53]) also independently conferred increased risk. Furthermore, a greater MetS burden progressively elevated DCM risk (*P*_trend_=0.026), with the adjusted HR increasing from 1.38 (1.05∼1.83) for 3 components to 2.35 (1.48∼3.72) for 5 components compared with 0 adverse components (Table 2).

Second, regarding MRS, we observed U-shaped associations of TG, SBP and DBP with DCM risk (Figure S1), whereas essentially linear relationships were observed for MRS, WC, HDL-C and HbA1c. Each 1-SD increase in MRS conferred a 25.9% (18.0∼34.3%) elevated risk of DCM, with a significant dose-response relationship across MRS quartiles (*P*_trend_=0.013). Notably, the effects of MRS and MetS were not comparable on distinct scales. Among the individual components, only WC (HR=1.36 [1.28∼1.45]) and HbA1c (HR=1.23 [1.16∼1.30]) were significantly associated with DCM risk (Table 2).

Third, even among participants without MetS, the MRS remained significantly associated with incident DCM risk (HR=1.25 [1.15∼1.34]). However, this association was absent in participants with ideal metabolic health (0 adverse MetS component; *P*=0.520), likely due to the limited statistical power from few DCM cases (*N*=72) in this subgroup.

### Robustness assessments of the MRS-DCM relationship

First, to evaluate the stability of the MRS calculated with equal weights, we recalculated this score via three distinct component-weighting schemes (Figure S2). The HRs of MRS calculated with positive WQS weights, normalized Cox effect sizes, and PC1 loadings were 1.31 (1.27∼1.35), 1.29 (1.25∼1.32), and 1.30 (1.22∼1.39) (Table S4), respectively, all of which aligned with the effects described above. Second, the MRS-DCM association remained stable after excluding the earliest incident cases and deaths, accounting for non-DCM mortality as a competing risk, or implementing GPS to balance confounder distributions (Table S5). Furthermore, the calculated E-values for significant traits (MRS: 1.83 [1.64∼2.02]; WC: 2.06 [1.88∼2.25]; HbA1c: 1.75 [1.59∼1.92]) exceeded the observed HRs (Table S5), suggesting that this association was resistant to potential residual confounding.

Notably, although no significant associations were observed for TG or DBP with DCM in the overall cohort, subgroup analyses revealed distinct patterns: TG was specifically related to DCM risk in younger individuals (HR=1.15 [1.02∼1.29]), whereas DBP was associated exclusively in females (HR=1.18 [1.05∼1.33]) (Table S6). Our time-stratified analysis demonstrated that MRS (short-term: HR=1.30 [1.19∼1.42]; long-term: 1.22 [1.11∼1.34]), WC (short-term: HR=1.37 [1.26∼1.50]; long-term: 1.35 [1.24∼1.47]), and HbA1c (short-term: HR=1.28 [1.18∼1.38]; long-term: 1.17 [1.08∼1.27]) exerted temporally robust influences on DCM, persisting across both short- and long-term intervals (Table S6).

### Contribution of individual MRS components in the MRS-DCM association

Given the presence of mild-to-moderate collinearity among the components of the MRS (|*r*|=0.02∼0.68; Figure 2A), we applied WQS regression to assess their relative contributions to incident DCM risk. Bidirectional WQS regression revealed a significant positive association between the weighted index of MRS components and DCM risk (RR=1.18, [1.12∼1.24]) but no significant negative association (*P*=0.88). The WQS analysis identified WC and HbA1c as the predominant contributors to this positive association, with weights of 0.58 and 0.22 (Figure 2B), respectively.

**Figure 2.**
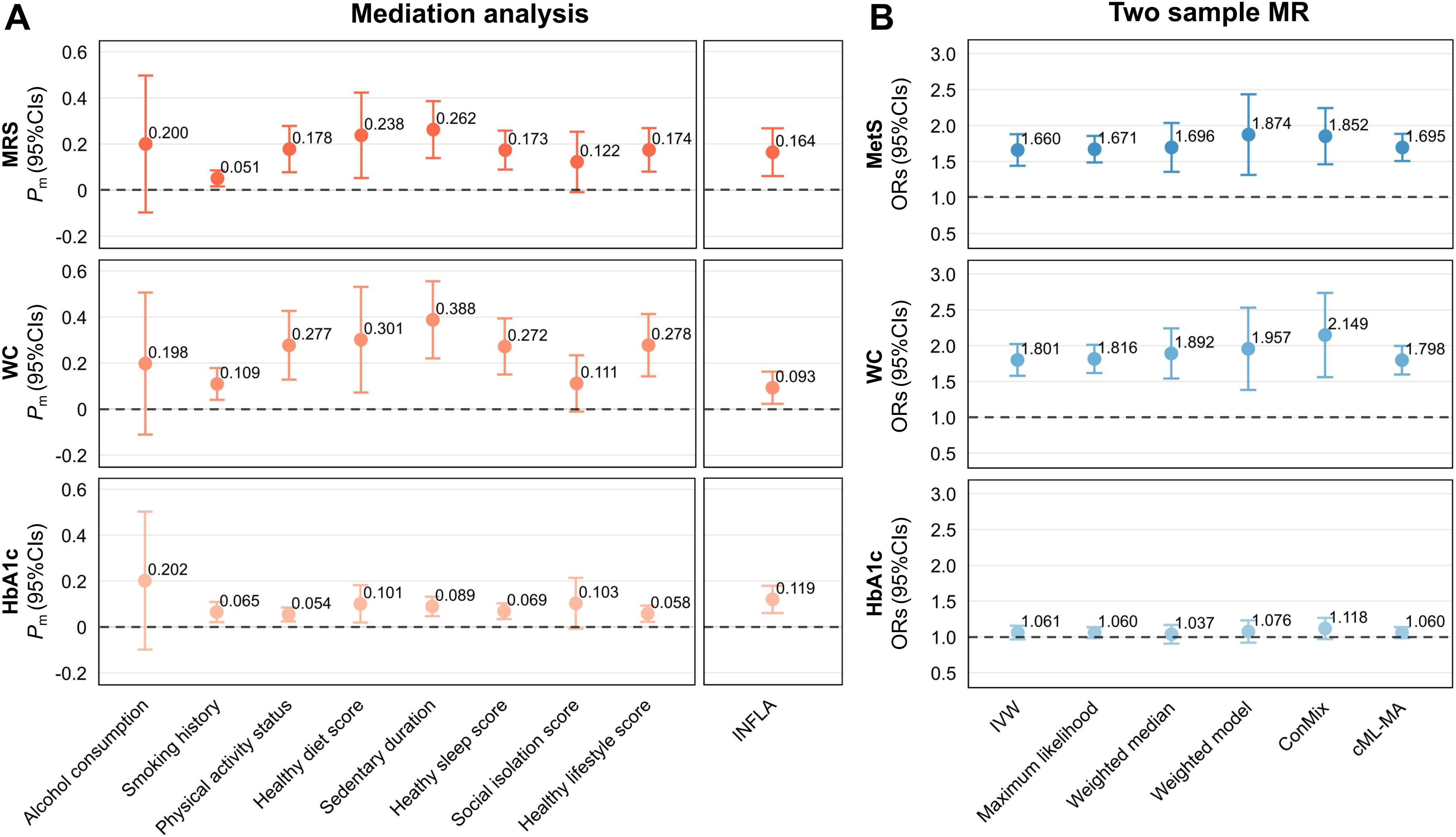
(A) The correlations between MRS components, expressed using Pearson’s correlation coefficients. (B) The proportional distribution of the weights of the positive and negative impacts of each component of MRS on relationship of MRS-DCM. (C) The temporal trends of MRS before the diagnosis of DCM. (D) The temporal trends of WC before the diagnosis of DCM. (E) The temporal trends of HbA1c before the diagnosis of DCM. MRS, metabolic risk score; DCM, dilated cardiomyopathy; WC, waist circumference; TG, triglycerides; HDL-C, high-density lipoprotein cholesterol; HbA1c, glycated hemoglobin; SBP, systolic blood pressure; DBP, diastolic blood pressure; RR, relative risk; CI, confidence interval.

### Temporal trends of MRS in DCM

Figure 2C-E depicts the longitudinal trajectories of MRS and its significant components (i.e., WC and HbA1c) over the 14 years preceding DCM diagnosis. During the period 12 to 14 years before diagnosis, no metabolic markers were significantly different between DCM patients and matched controls. Divergent trajectories emerged thereafter: WC exceeded those of controls starting at 12 years pre-diagnosis, followed by MRS divergence at 10 years and HbA1c at 9 years. HbA1c exhibited a monotonic upwards trend in DCM patients as the diagnosis progressed. In contrast, the MRS and WC plateaued at 7 years pre-diagnosis before gradually declining to levels near those of the control group.

### Mediating path from the lifestyle-MRS-DCM and MRS-inflammation-DCM

As shown in Figure 3A and Table S7, the MRS partially mediated the associations of several lifestyle factors, including healthy diet score, physical activity status, sedentary duration, healthy sleep score, and the composite healthy lifestyle score, with incident DCM risk. The *P_m_* by MRS ranged from 5.1% (1.6∼8.5%) for smoking history to 26.2% (13.9∼38.6%) for sedentary duration. Among the metabolic components comprising MRS, WC exhibited the strongest mediation effects, with *P_m_* reaching 10.9% (4.0∼17.8%) for smoking history and 38.8% (22.0∼55.5%) for sedentary duration. In contrast, HbA1c displayed weaker mediation, with *P_m_* ranging from 5.4% (2.4∼8.5%) for physical activity status to 10.1% (1.9∼18.2%) for healthy diet score. We also detected a significant mediating role of INFLA in the associations of MRS (*P_m_*_□_=□16.4 [6.1∼26.8]), WC (*P_m_*=c9.3 [2.3∼16.3]), and HbA1c (*P_m_*=11.9 [6.0∼17.9]) with DCM risk (Figure 3A and Table S8).

**Figure 3.**
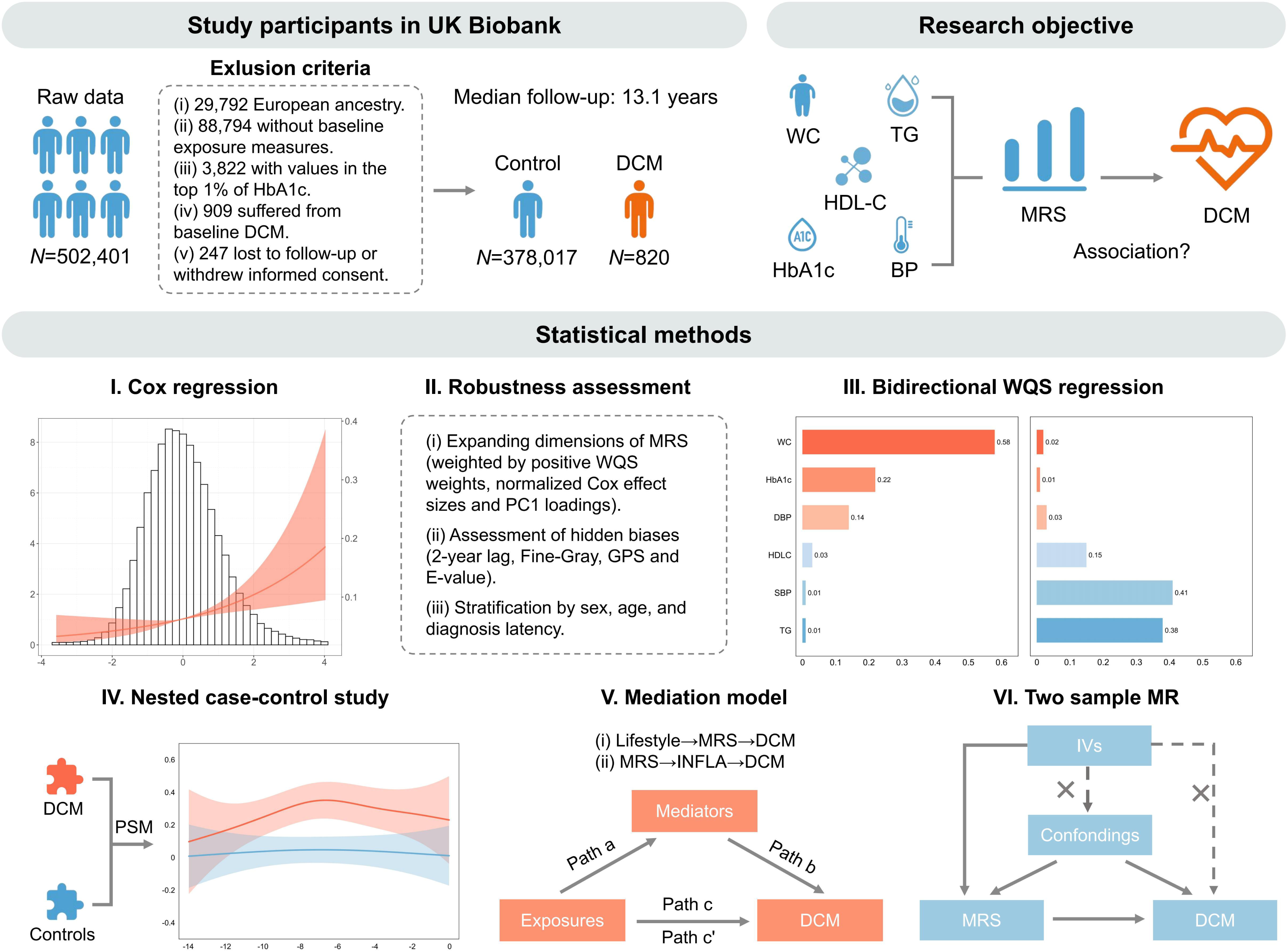
(A) The mediated proportions of MRS/its key components in the association between lifestyle and DCM, and the mediated proportion of INFLA in the association between MRS/its key components and DCM. (B) Forest plot of MR for the assessment of causality between MetS and DCM. MRS, metabolic risk score; WC, waist circumference; HbA1c, glycated hemoglobin; MetS, metabolic syndrome; DCM, dilated cardiomyopathy; INFLA, low-grade inflammation score; MR, Mendelian Randomization; IVW, inverse variance weighted method; ConMix, contamination mixture method; cML-MA, constrained maximum likelihood and model averaging method; *P*_m_, mediation proportion; OR, odds ratio; CI, confidence interval.

### Genetic causal association between MetS and DCM

Finally, we employed MR to assess causality from a genetic perspective. The PVEs by the IVs (Tables S9-S11) of MetS (*N*_IV_=520), WC (*N*_IV_=351) and HbA1c (*N*_IV_=340) were 2.8%, 4.4% and 11.0%, respectively. All *F*-statistics (29.8∼1695.0) substantially exceeded the conventional threshold of 10, mitigating weak instrument bias. Appropriate IVW estimates were selected via the Cochran’s *<u>Q</u>* test (Table S12) and validated via five additional MR methods. Genetically predicted MetS (OR_IVW_=1.65 [1.45∼1.88]) and WC (OR_IVW_=1.79 [1.58∼2.03]) were significantly causally associated with increased DCM risk. However, no causal association was observed for HbA1c (*P*>0.05; Figure 3B and Table S13). MR-Egger intercept tests indicated no significant horizontal pleiotropy for the observed causal associations (Table S14). The results remained robust after excluding outliers identified via MR-PRESSO (Table S14). Leave-one-out sensitivity analyses also confirmed that there were no influential individual SNPs (Figure S3), and Steiger directionality tests supported the causal direction (Table S14).

## Discussion

### Restatement of the core findings

Using a multi-model evidence framework, our study revealed a significant relationship between MRS and DCM incidence, persisting even among non-MetS individuals. Robustness analyses confirmed stable results under variations in weighting schemes, confounding factors, and time intervals. WQS regression identified abdominal obesity and hyperglycemia as key contributors to this link. Temporal analyses revealed that DCM patients presented elevated MRS trajectories beginning approximately one decade prior to diagnosis. Mediation analyses further implicated MRS as a partial intermediary linking lifestyle factors to DCM, with INFLA mediating the MRS-DCM link. MR analyses provided causal evidence that genetically determined MetS increases the risk of DCM development.

### Engagement with prior research

#### Extending the MetS-DCM paradigm

Previous studies have explored the role of metabolic dysfunction in the pathogenesis of the myocardium 3,33. For example, a cohort study indicated an association between MetS and heart failure risk, with a HR of 1.32 (1.16∼1.51) 34; Nyman et al. further validated MetS-driven left ventricular dysfunction via cardiac magnetic resonance imaging radiomics 35. Using the sufficiently representative UK Biobank and rigorous multi-model causal frameworks, our study elucidated the association between MRS and DCM for the first time, thereby significantly consolidating and extending the metabolic-heart failure paradigm.

Our finding that metabolic dysfunction precedes and predicts DCM incidence provides a crucial etiological link between two seemingly disparate strands of prior research. It reconciles the mechanistic evidence of MetS-induced myocardial injury 6 with observations of its complex clinical prognosis 5. We demonstrate that these phenomena are not contradictory but sequential: MetS is a root cause of DCM, whose detrimental cardiac effects can later be obscured in advanced stages by confounding factors like the obesity paradox. Our work thus repositions metabolic health from a mere disease modifier to a fundamental element in DCM pathogenesis.

#### Strong, ambiguous, or heterogeneity effects between components and the DCM

Among MRS components, WC showed the strongest association with incident DCM (HR=1.36 [1.28∼1.45]), reinforcing the cardiovascular salience of abdominal adiposity and aligning with population evidence that excess adiposity tracks with myocardial remodelling and failure 36. Corroborating Antonopoulos and colleagues’ findings 37, an analysis of the 2015-2019 U.S. National Inpatient Sample revealed that hospitalized patients with a body mass index ≥30 kg/m² presented an 82% (79∼84%) greater risk of DCM than did normal-weight individuals.

In parallel, HbA1c was demonstrated to be positively associated with DCM in our cohort, a finding that complements prior observational work linking glycemic burden to impaired ventricular function 38 and subclinical myocardial dysfunction 39. However, genetic triangulation urges caution for glucose metabolism: our MR analysis did not support a causal HbA1c-DCM effect, unlike the consistent signals for MRS and adiposity. This divergence may reflect underlying differing exposure windows, residual confounding, and instrument limitations 40.

Blood pressure illustrates another interesting finding. Although DBP showed no overall association, it was significantly linked to risk in women. Sex-specific myocardial energetics, increased estrogen-modulated fatty-acid oxidation, and reduced diastolic perfusion due to elevated DBP offer a mechanistic basis for this sex difference 41. Moreover, lipids underscore life-stage heterogeneity: TG was null overall yet predicted DCM in younger participants, plausibly via a hormonal milieu that amplifies *PPAR-*γ signalling and adipokine (e.g., leptin) release, promoting myocardial fibrosis through *STAT3* 42. Finally, the binary classification of HDL-C (whether HDL-C is reduced or not) displayed a significant correlation with the risk of DCM, differing from the performance of HDL-C as a continuous variable 38. This threshold effect suggests that the protective role of HDL-C in DCM may depend on its functional integrity rather than its absolute concentration alone 43.

#### Validity of the refined score and weighting schemes

Owing to the lack of fasting insulin in the UK Biobank, we used HbA1c instead of homeostasis model assessment of insulin resistance (HOMA-IR) when calculating the MRS 10,16. While HOMA-IR better captures early insulin resistance, HbA1c offers superior standardization and stability and reflects chronic (8∼12 weeks) glycemic exposure 44, aligning with DCM pathophysiology 45. Furthermore, despite reduced sensitivity to some acute diseases (e.g., acute dysglycemia, acute anaemia) 46, HbA1c significantly contributes to DCM risk (HR=1.23; 22% of WQS weight) and shows a progressive increase in pre-diagnosis. Therefore, we considered it appropriate to employ HbA1c in our analyses where DCM is the primary outcome.

In particular, weighted MRSs were demonstrated to be consistently related to DCM risk, with effect sizes comparable with those of the unweighted score, indicating that weighting refined the emphasis of individual components (e.g., elevating contributions for WC and HbA1c) without altering our primary conclusions. This robustness likely stems from the DCM’s chronic drivers (central adiposity, hyperglycemia) dominating risk. Generalizability to other contexts requires more external validation, as optimal weights may differ for diseases driven by acute lipidomic or hemodynamic stressors 47.

### Elucidation of possible biological mechanisms

#### Obesity-driven metabolic-inflammatory cascade in DCM

Our study implicates central adiposity and hyperglycaemia as key drivers of DCM pathogenesis. Abdominal obesity, quantified by elevated WC, reflects aberrant adipokine signalling and directly impairs cardiac function via lipotoxicity and altered substrate utilization. Experimental and clinical evidence indicates that obesity enhances myocardial fatty acid uptake, which, coupled with mitochondrial uncoupling protein upregulation, reduces adenosine triphosphate synthesis efficiency 48. Concomitant ceramide accumulation and lipotoxic intermediates trigger cardiomyocyte apoptosis and contractile dysfunction 49. Moreover, our mediation analyses indicate that this metabolic state drives chronic low-grade inflammation. This oxidatively triggered inflammation may propagate myocardial injury through cardiomyocyte apoptosis, fibrosis, and diastolic dysfunction, culminating in DCM 45.

#### Hyperglycemia-induced advanced glycation end product accumulation and insulin resistance synergistically impair myocardial function

In terms of glucose, chronic hyperglycemia accelerates advanced glycation end-product (AGE) accumulation. These compounds bind myocardial AGE receptors, inducing oxidative stress, inflammatory signalling, and TGF-β-mediated fibrosis, collectively driving cardiac structural remodelling 50. Simultaneously, elevated HbA1c exacerbates cardiomyocyte insulin resistance, impairing glucose utilization and redirecting metabolism toward fatty acid oxidation. This shift promotes cytotoxic lipid deposition and ceramide-induced mitochondrial dysfunction, synergizing with glycative stress to worsen ectopic fat accumulation 51. Critically, these insults disrupt calcium handling through sarcoplasmic reticulum dysfunction. Finally, the resulting calcium dyshomeostasis manifests as systolic impairment and ventricular dilation, which are characteristic of DCM pathology 52.

### Public health significance

The established association between the MRS and DCM provides novel insights for public health strategies. First, metabolic trajectory management should become a key strategy for DCM prevention and control. DCM patients exhibited abnormal increases in MRS, WC, and HbA1c approximately 10 years before diagnosis, suggesting that clinical screening windows need to be significantly advanced. We recommend constructing a DCM risk prediction model incorporating WC and HbA1c. Second, lifestyle interventions should target modifiable metabolic intermediaries. Community health programs should prioritize WC and glucose control by addressing behavioural risks such as poor diet, physical inactivity, sedentary lifestyles, and poor sleep quality.

For risk-stratified management, we advocate for a metabolic phenotype screening system. High-risk individuals with abdominal obesity and glucose metabolism abnormalities should undergo cardiac ultrasound and biomarker testing at shorter intervals (e.g., every 2 years), whereas patients with central adiposity and hyperglycemia may extend follow-up intervals to longer periods (e.g., every 7 years). Public health organizations ought to update their guidelines so as to incorporate dynamic MRS evaluations into DCM prevention plans. Moreover, as the benefits offered by drugs like GLP-1 receptor agonists to visceral fat and myocardial protection are obvious 53, it is advisable for healthcare policies to grant preference to medications for those who, despite persistent elevated WCs and behavioural change unresponsiveness, still struggle and could be a turning point in reducing the occurrence rate of DCM.

### Strengths of our research

This study overcomes the limitations of the traditional dichotomous diagnosis of MetS 54, which adopts a continuous metabolic scoring system 10. By standardizing each component through the *Z*-score and integrating them with component-specific or even equal weights, this scoring system overcomes the neglect of the degree of abnormality and synergistic effects by the traditional “3 out of 5” rule. It enables the quantification of the cumulative burden of metabolic disorders and sensitively captures the DCM risk of individuals with borderline abnormal values (such as those whose WC is only 1 cm less than the threshold). In addition, this system adjusts for factors such as age, gender, and height as corresponding indicators, effectively controlling for individual differences.

Another advantage is that our study establishes a multi-model causal framework that integrates survival analysis, WQS regression, metabolic trajectory modelling, and MR. This approach uniquely captures dynamic pre-diagnostic dysregulation patterns through serial metabolic assessments in a 14-year nested case-control cohort.

Critically, comprehensive sensitivity analyses exceed conventional standards, including competing risk adjustments, E-value quantification for unmeasured confounding, generalized propensity scoring for selection bias, and weight calculation robustness testing. The mediation framework unveils metabolic dysregulation as the central conduit linking lifestyle to DCM pathogenesis, substantiated by rigorous pathway validation. Bidirectional MR further minimizes reverse causation via Steiger testing and pleiotropy-robust estimators.

### Limitations of our research

This study has several limitations. First, MRS and its components were measured only at baseline. Longitudinal changes in metabolic parameters during follow-up were not captured, potentially underestimating the dynamic nature of metabolic dysfunction. Second, the absence of omics data such as myocardial biopsy or circulating biomarkers and the inability to fully elucidate the molecular bridge between metabolic components and myocardial injury are also problems that we cannot solve for the time being. Third, MR analysis’s reliance on multivariate GWAS data of MetS 18 creates a definition with our continuous metabolic scoring system employed in observational analysis, potentially precluding mutual result corroboration.

### Conclusions

Our study establishes metabolic dysfunction as a pivotal, modifiable driver of DCM. The robust, causal link between metabolic risk factors (especially central adiposity and hyperglycemia) and DCM underscores the paramount importance of prioritizing metabolic health for primary prevention. Additionally, our findings reveal that metabolic abnormalities manifest approximately a decade prior to DCM diagnosis, providing a crucial window for intervention. Crucially, we identified two key mechanistic pathways: metabolic risk factors significantly mediate the detrimental impact of adverse lifestyles on DCM, whereas inflammation accounts for a substantial portion of the metabolic risk effect.

## List of abbreviations

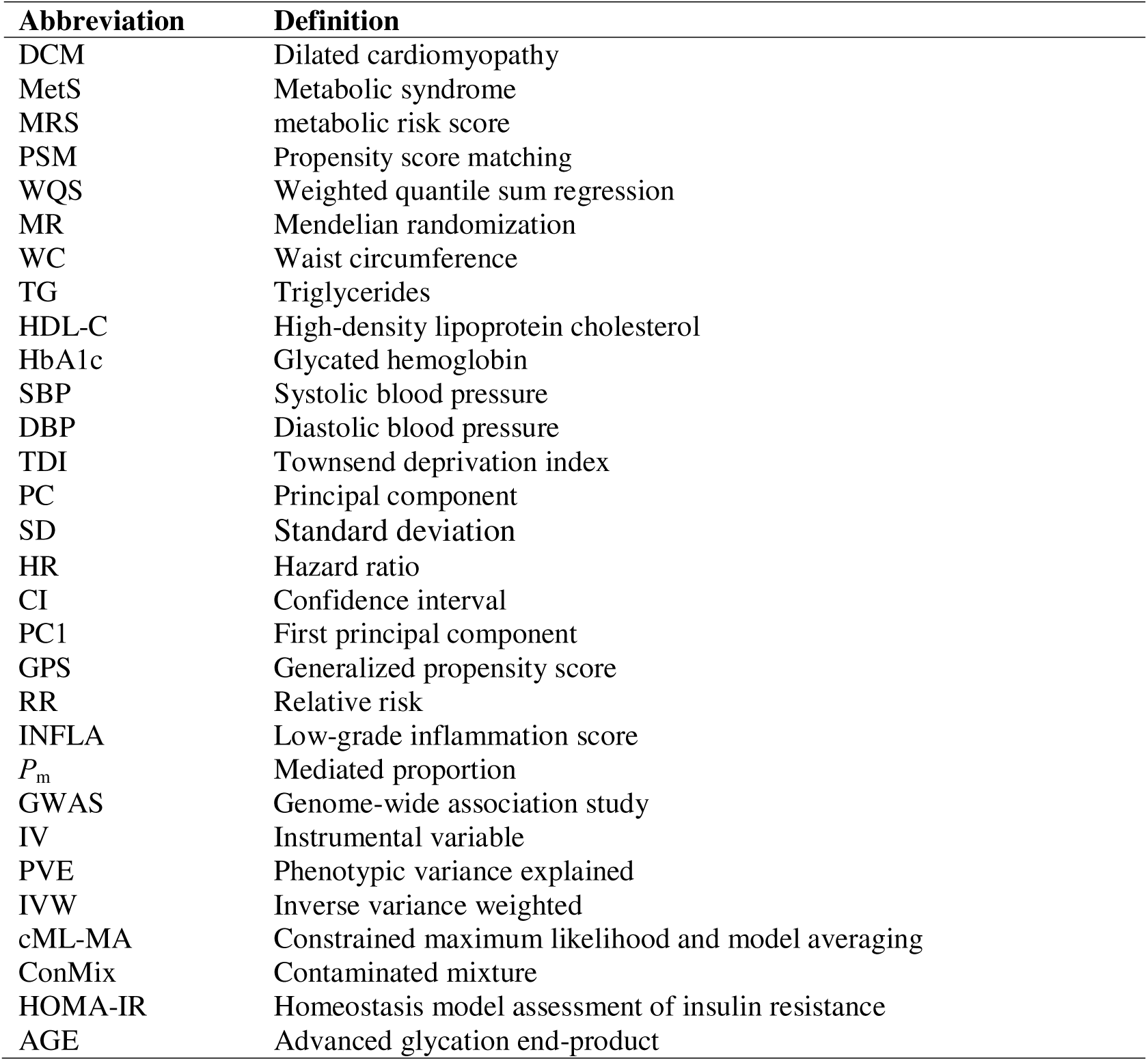

| Abbreviation | Definition |
| --- | --- |
| DCM | Dilated cardiomyopathy |
| MetS | Metabolic syndrome |
| MRS | metabolic risk score |
| PSM | Propensity score matching |
| WQS | Weighted quantile sum regression |
| MR | Mendelian randomization |
| WC | Waist circumference |
| TG | Triglycerides |
| HDL-C | High-density lipoprotein cholesterol |
| HbA1c | Glycated hemoglobin |
| SBP | Systolic blood pressure |
| DBP | Diastolic blood pressure |
| TDI | Townsend deprivation index |
| PC | Principal component |
| SD | Standard deviation |
| HR | Hazard ratio |
| CI | Confidence interval |
| PC1 | First principal component |
| GPS | Generalized propensity score |
| RR | Relative risk |
| INFLA | Low-grade inflammation score |
| $P_m$ | Mediated proportion |
| GWAS | Genome-wide association study |
| IV | Instrumental variable |
| PVE | Phenotypic variance explained |
| IVW | Inverse variance weighted |
| cML-MA | Constrained maximum likelihood and model averaging |
| ConMix | Contaminated mixture |
| HOMA-IR | Homeostasis model assessment of insulin resistance |
| AGE | Advanced glycation end-product |

## Additional File

Supplementary Methods, Supplementary Figures and Supplementary Tables.

## Acknowledgements

This study was based on the UK Biobank resource under application number 88159. The UK Biobank was established by the Wellcome Trust Medical Charity, Medical Research Council, Department of Health, Scottish Government, and Northwest Regional Development Agency. It has also received funding from the Welsh Assembly Government, the British Heart Foundation and Diabetes UK. We also thank all the GWAS consortia for making summary statistics publicly available for us and are grateful to all the investigators and participants who contributed to those studies. The data analyses in the present study were carried out with a high-performance computing cluster that was supported by the special central finance project of local universities for Xuzhou Medical University.

## Authors’ contributions

PZ conceived the idea for the study. PZ obtained the data. PZ and JQ cleared the datasets; PZ and JQ performed the data analyses. PZ, WS, JQ, SZ, YJ, HL, YX, CZ, YY, KW and HC interpreted the data analysis results. PZ and JQ wrote the manuscript with help from the other authors.

## Ethics approval and consent to participate

Not applicable.

## Availability of data and materials

The data supporting this study are from two primary sources. Individual-level data were obtained from the UK Biobank under Application ID 88159, which researchers can access by applying through the official website (https://www.ukbiobank.ac.uk/). Additionally, this study utilized summary-level data from the GWAS Catalog (https://www.ebi.ac.uk/gwas). All data generated from these sources or analyzed during the current study are included in this article and its supplementary files.

## Data availability statement

No new data were generated or analysed in support of this research.

---

Table 1.Baseline characteristics of all included participants in the UK Biobank cohort.

Notes: The differences in characteristics between incident DCM cases and non-DCM participants were evaluated by two-sample t tests or Pearson’s χ2 tests. DCM, dilated cardiomyopathy; MRS, metabolic risk score; WC, waist circumference; TG, triglyceride; HDL-C, high-density lipoprotein cholesterol; HbA1c, glycated hemoglobin; SBP, systolic blood pressure; DBP, diastolic blood pressure; TDI, Townsend deprivation index; N, sample size.

Table 2. Phenotypic associations between MRS, MetS and DCM.

Note: MRS, metabolic risk score; DCM, dilated cardiomyopathy; HR, hazard ratio; CI, confidence interval; WC, waist circumference; TG, triglycerides; HDL-C, high-density lipoprotein cholesterol; HbA1c, glycated hemoglobin; SBP, systolic blood pressure; DBP, diastolic blood pressure; MetS, metabolic syndrome; HPT, hypertension; Ref., reference group.

