## Supplementary Figures for "An integrative study of risk assessment, mediation analysis, and causal inference for the relationship between metabolic syndrome and dilated cardiomyopathy"

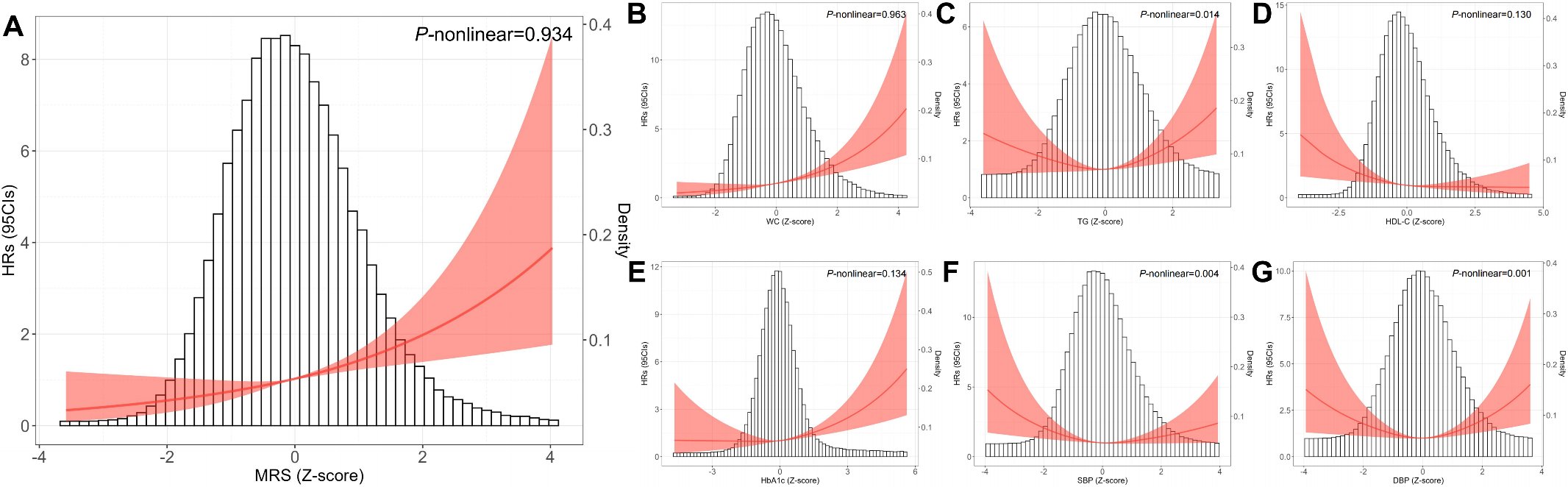

Figure S1. (A) Restricted cubic spline curve of the MRS-DCM relationship. (B) Restricted cubic spline curve of the WC-DCM relationship. (C) Restricted cubic spline curve of the TG-DCM relationship. (D) Restricted cubic spline curve of the HDL-C-DCM relationship. (E) Restricted cubic spline curve of the HbA1c-DCM relationship. (F) Restricted cubic spline curve of the SBP-DCM relationship. (G) Restricted cubic spline curve of the DBP-DCM relationship. MRS, metabolic risk score; WC, waist circumference; TG, triglycerides; HDL-C, high-density lipoprotein cholesterol; HbA1c, glycated hemoglobin; SBP, systolic blood pressure; DBP, diastolic blood pressures; DCM, dilated cardiomyopathy; HR, hazard ratio; CI, confidential interval.

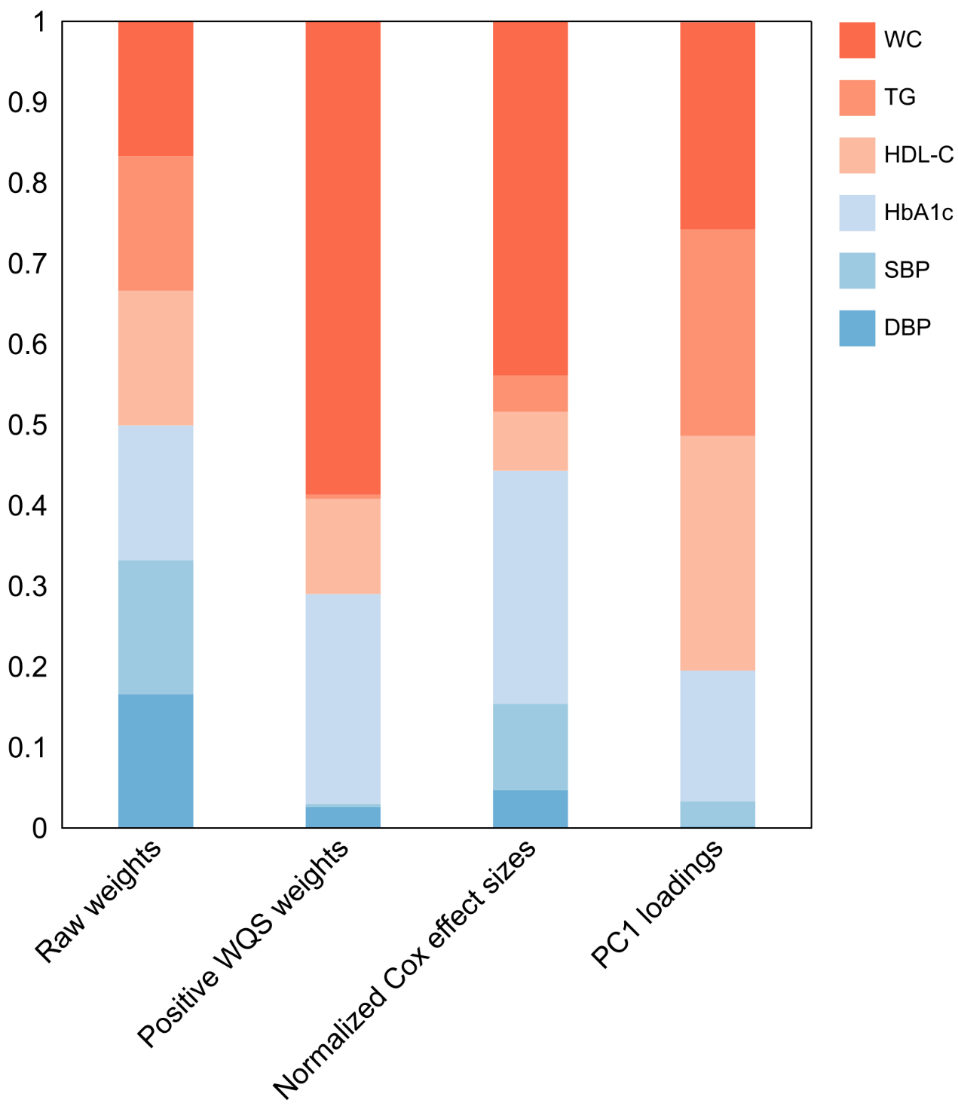

Figure S2. Weights assigned to MRS's components, derived from primary model and three component-weighted schemes. MRS, metabolic risk score; WC, waist circumference; TG, triglycerides; HDL-C, high-density lipoprotein cholesterol; HbA1c, glycated hemoglobin; SBP, systolic blood pressure; DBP, diastolic blood pressures; WQS, weighted quantile sum; PCA, principal component analysis.

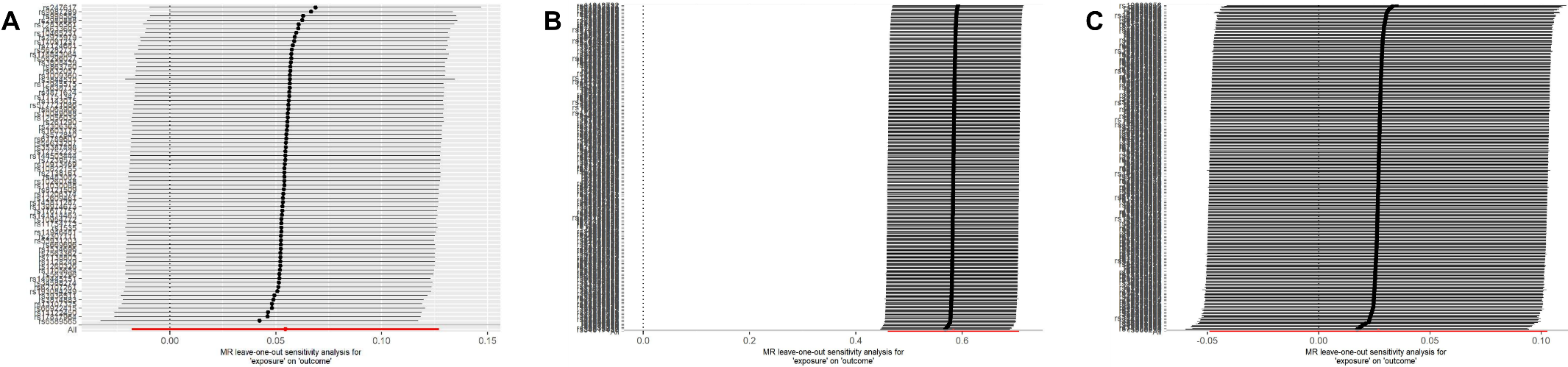

Figure S3. (A) Individual SNP contributions of the causal association between genetically predicted MetS and DCM. (B) Individual SNP contributions of the causal association between genetically predicted WC and DCM. (C) Individual SNP contributions of the causal association between genetically predicted HbA1c and DCM. MetS, metabolic syndrome; DCM, dilated cardiomyopathy; WC, waist circumference; HbA1c, glycated hemoglobin.
