## Supplementary Methods for "An integrative study of risk assessment, mediation analysis, and causal inference for the relationship between metabolic syndrome and dilated cardiomyopathy"

**Method S1. Treatment for covariates**

We categorized these covariates into three groups: sociodemographic characteristics and lifestyle factors. Sociodemographic characteristics include age (field 21022, continuous), gender (field 31, male or female), educational level (field 6138, with or without a university degree), income (fields 738, <£18,000, £18,000~£30,999, £31,000~£51,999, £52,000~£100,000, or >£100,000) and the Thompson deprivation index (TDI, field 189). Lifestyle factors were obtained from self-reports and included smoking history (fields 1249 and 20160, previous smoking or not) and alcohol consumption history (fields 3731 and 20117, previous alcohol consumption or not). Physical activity status was assessed via the International Physical Activity Questionnaire (IPAQ) short form 1. Physical activity (field 22032) was categorized into three groups on the basis of this questionnaire: low (<600 minutes/week), moderate (600-3000 minutes/week), and high (>3000 minutes/week). Additionally, a healthy diet score was calculated on the basis of food consumption frequency 2. One point was assigned for each of the following favorable conditions: (i) vegetable intake (fields 1289 and 1299) ≥ four tablespoons per day; (ii) fruit intake (fields 1309 and 1319) ≥ three pieces per day; (iii) fish intake (fields 1329 and 1339) ≥ twice per week; (iv) unprocessed red meat intake (fields 1369, 1379 and 1389) ≤ twice per week; and (v) processed meat intake (field 1349) ≤ twice per week. The cumulative scores of each condition ranged from 0 to 5, representing the overall scores of a healthy diet. Additionally, we included the values of the top ten principal components (field 22009) as covariates.

Educational qualifications were unavailable for 67,588 participants (17.8%), and household income before tax was missing for 49,827 participants (13.2%). Information on physical activity was missing for 71,300 participants (18.8%), whereas healthy diet score data were absent for 55,725 participants (14.7%). In addition, the TDI was missing for 448 participants (0.1%).

**Method S2. Quality control of GWAS summary-level data**

Following previous studies 3^,^4, we performed stringent quality control procedures on the used genome-wide association study summary-level datasets as follows: (i) duplicated single nucleotide polymorphisms (SNPs) were removed; (ii) the data were restricted to biallelic SNPs; (iii) SNPs without rs labels were excluded; (iv) SNPs that were not genotyped in the 1000 Genomes Project Phase 3 or whose alleles did not match those that were removed; (v) the major histocompatibility complex region (chr6: 28.5-33.5Mb); (vi) SNPs with a minor allele frequency >0.01 were retained.

**Method S3. Treatment for binary metabolic syndrome (MetS)**

MetS was defined based on a joint statement by the American Heart Association/National Heart, Lung, and Blood Institute (AHA/NHLBI) in 2009 5, as the presence of at least three of the following five components: (i) elevated waist circumference (WC; ≥102 cm in males and ≥88 cm in females); (ii) elevated triglycerides (TG; ≥150 mg/dL or 1.7 mmol/L); (iii) reduced high-density lipoprotein cholesterol (HDL-C; <40 mg/dL or 1.0 mmol/L in males; <50 mg/dL or 1.3 mmol/L in females); (iv) elevated fasting glucose (≥100 mg/dL); (v) elevated blood pressure or hypertension (systolic blood pressure [SBP] ≥130 mmHg or diastolic blood pressure [DBP] ≥85 mmHg). Considering the lack of fasting blood glucose data, we used the more stable glycated hemoglobin (HbA1c; >42.0 mmol/mol can be dedined as elevated HbA1c) measurement according to the WHO/IDF guidelines instead 6^,^7. Additionally, we excluded participants in the top 1% of HbA1c to address potential bias from extreme values in HbA1c distributions.

**Method S4. Construction of component-weighted metabolic risk scores (MRSs) and validation**

We began with 6 MetS components (WC, TG, HDL-C, HbA1c, SBP, and DBP), measured at baseline. All composite scores were constructed as weighted sums

$$w_{\text{WC}}\text{Z}_{\text{WC}}\text{+(}w_{\text{TG}}\text{Z}_{\mathrm{TG}}\text{-}w_{\text{HDL-C}}\text{Z}_{\text{HDL-C}}\text{)/2+}w_{\text{HbA1c}}\text{Z}_{\text{HbA1c}}\text{+(}w_{\text{SBP}}\text{Z}_{\text{SBP}}\text{+}\text{w}_{\text{DBP}}\text{Z}_{\text{DBP}}\text{)/2}$$

We considered three versions of weighted MRS that differed only by how *w* was obtained.

(1) *Positive weighted quantile sum (WQS) weights (supervised).* Each component was quantile-coded to *Q_j_*. A WQS index was estimated under a single risk-increasing direction (positive) constraint and used to predict incident DCM

$$\mathrm{WQS}=\sum_{j=1}^{6} w_{j}Q_{j}$$

All these weights satisfy the following constraints

$w_{j}\geq0$ and$\sum_{j=1}^{6} w_{j}=1$

(2) *Normalized Cox effect sizes (supervised).* We fit a multivariable Cox model for DCM including all components and extracted the effect size $\beta_{j}^{*}$. The weights were defined as the absolute effect szies, normalized to sum to one

$$w_{j}=\frac{\left| \beta_{j}^{*} \right|}{\sum_{j=1}^{6} \left| \beta_{j}^{*} \right|}$$

(3) *First principal component (PC1) loadings (unsupervised).* We performed principal component analysis on the components' correlation matrix and took the PC1 loadings as weights.

*Monte Carlo cross-validation for supervised weights.* For WQS and Cox-derived weights, we generated 1,000 random 50%/50% training-validation splits. In each split we: (i) estimated weights in the training set; (ii) constructed the MRS in the held-out validation set; (iii) fit a Cox model for DCM with the MRS as the exposure with the same covariate adjustment as the main analysis. HRs were aggregated as means with 95%CIs across all iterations.

*Evaluation of the unsupervised PC1 score.* Because PC1 weights did not use outcome information, they were computed on the full dataset; association with incident DCM was then assessed via Cox regression with the same set of covariates.

**Method S5. Treatment for lifestyle variables, healthy** **lifestyle score and low-grade inflammation score (INFLA)**

The assessment methods for the four routine factors, including healthy diet score, alcohol consumption history, smoking history, and physical activity status, are consistent with those used for the covariates. The sedentary duration 8 is the sum of the daily TV-watching (field 1070) and computer-using (field 1080) times. The healthy sleep score 9 is constructed by integrating five sleep-related components, with each component accounting for one point of the total score: (i) sleep duration (field 1160, 7-8 hours per day); (ii) morning/evening person (field 1180, "morning" or "more morning than evening"); (iii) insomnia (field 1200, "never/rarely" or "sometimes"); (iv) no snoring (field 1210); and (v) daytime dozing/sleeping (field 1220, "never/rarely" or "sometimes"). The total score ranges from 0 to 5, with a higher value indicating a healthier sleep pattern. The social isolation score 10 is calculated on the basis of the sum of the following three indices, with each component accounting for one point of the total score: (i) the frequency of friends/family visits (field 1031, "once a month", "a few months", "never", "almost never", "no friends" or "family outside the household"); (ii) participation in leisure/social activities (field 6160, "none of the above"); and (iii) living alone or not (field 709, "living alone"). The total score ranges from 0 to 3, with a higher value indicating a higher level of social isolation. Ultimately, we assigned one point for each of the following: a healthy diet score of 4 or above, a history of smoking, a history of alcohol consumption, moderate or high levels of physical activity, sedentary duration of less than 4 hours, a healthy sleep score of 4 or above, and a social isolation score of less than 1. The total score ranges from 0 to 7, with a higher score indicating a more regular and healthier lifestyle 8.

The INFLA score 11 integrates four synergistic inflammatory markers, including C-reactive protein (field 30710), white blood cell count (field 30000), platelet count (field 30080), and neutrophil-lymphocyte ratio (fields 30140 and 30120), and all components in the highest deciles (7th to 10th) are assigned values from +1 to +4, whereas biomarker levels in the lowest deciles (1st to 4th) are assigned values from -4 to -1. INFLA scores range from -16 to +16, with higher scores indicating higher levels of low-grade inflammation 12. In addition to the INFLA score, all inflammatory markers were converted to natural logarithmic values and standardized prior to analysis.

**Method S6. Modelling assumptions for Mendelian randomization**

To perform valid causal inference in Mendelian randomization (MR) analysis, the SNPs used as instrumental variables (IVs) must satisfy three key assumptions 13^,^14. These include the relevance assumption, which requires that IVs are strongly associated with the exposure; the independence assumption, which states that IVs must not be related to any confounders; and the exclusion restriction assumption, which necessitates that IVs are associated with the outcome solely through the exposure pathway and do not exhibit any pleiotropic effects.

We performed the following steps to satisfy the relevance assumption: we applied the clumping procedure of PLINK 15 by setting the significance level, LD, and physical distance to 5×10^-8^, 0.01 and 10000 kb, respectively, with genotypes of 503 individuals from the 1000 Genomes Project as the reference panel 16. Phenotypic variance explained (PVE) by IVs and the *F*-statistic were employed to evaluate the strength of IVs, with an *F*-statistic ≥10 indicating that the IV is strongly associated with the exposure 17^,^18. Briefly, PVE=$\hat{\beta}^{2}$/($\hat{\beta}^{2}$+*se*^2^×*N*), *F*=PVE×(*N*-2)/(1-PVE), where $\hat{\beta}$ is the effect size of MetS and its components, *se* is the standard error of the effect size, and *N* is the effective sample size.

To ensure that the independence assumption and exclusion restriction assumption were satisfied, we undertook a series of rigorous procedures. We excluded IVs that were significantly associated with potential confounders, using a stringent threshold of *P*<5×10^-8^, including factors such as BMI 19, education 20, income 21, smoking initiation and alcohol consumption 22. This approach ensured that the IVs were not linked to confounders that could bias the exposure-outcome relationship. Additionally, we removed IVs that were likely related to the outcome, applying a conservative Bonferroni-corrected threshold of *P*<0.05/number of SNPs, to protect against horizontal pleiotropy, where IVs could influence the outcome through pathways other than the exposure.

We estimated causal associations primarily via the inverse variance weighted method 23, which assumes that all instruments are valid, and additionally applied five methods 24 to reduce false positives. The maximum likelihood method 25 handles uncertainty in SNP-exposure associations and adjusts for sample overlap via a correlation parameter (psi). The weighted median 26 and weighted mode 27 methods mitigate the influence of outlier instruments by using the median and the most frequent estimate, respectively. The contamination mixture method 28 constructs a likelihood function from individual causal estimates, providing robust results even when many instruments are invalid. Finally, the constrained maximum likelihood and model averaging methods 29 address both related and unrelated pleiotropy to exclude invalid instruments and yield reliable causal estimates.
